# Effectiveness of digital interventions to improve household and community infection prevention and control behaviours and to reduce incidence of respiratory and/or gastro-intestinal infections: A rapid systematic review

**DOI:** 10.1101/2020.09.07.20164947

**Authors:** Natalie Gold, Xiao-Yang Hu, Sarah Denford, Ru-Yu Xia, Lauren Towler, Julia Groot, Rachel Gledhill, Merlin Willcox, Ben Ainsworth, Sascha Miller, James Denison-Day, Michael Moore, Cathy Rice, Jennifer Bostock, Beth Stuart, Kate Morton, Paul Little, Richard Amlôt, Tim Chadborn, Lucy Yardley

**Affiliations:** Public Health England Behavioural Insights, Public Health England, UK; Faculty of Philosophy, University of Oxford, UK; Primary Care Population Sciences and Medical Education, University of Southampton, UK; Bristol Medical School (PHS), Faculty of Health Sciences, University of Bristol; Centre for Evidence-Based Chinese Medicine, Beijing University of Chinese Medicine, China; School of Psychology, University of Southampton, UK; Department of Psychology, University of Bath, UK; Public Contributor; Policy Research Unit, London School of Hygiene & Tropical Medicine, UK; School of Psychological Science, University of Bristol, UK

**Author notes:** Corresponding author: Natalie Gold.

**Keywords:** behaviour, behaviour change, behavioural medicine, community health, COVID-19, digital medicine, eHealth, hygiene, handwashing, infection control, novel coronavirus

## Abstract

**Background:** Digital interventions have potential to efficiently support improved hygiene practices to reduce transmission of COVID-19.

**Objective:** To evaluate the evidence for digital interventions to improve hygiene practices within the community.

**Methods:** We reviewed articles published between 01 January 2000 and 26 May 2019 that presented a controlled trial of a digital intervention to improve hygiene behaviours in the community. We searched MEDLINE, Embase, PsycINFO, Cochrane Controlled Register of Trials (CENTRAL), China National Knowledge Infrastructure and grey literature. Trials in hospitals were excluded, as were trials aiming at prevention of sexually transmitted infections; only target diseases with transmission mechanisms similar to COVID-19 (e.g. respiratory and gastrointestinal infections) were included. Trials had to evaluate a uniquely digital component of an intervention. Study designs were limited to randomised controlled trials, controlled before-and-after trials, and interrupted time series analyses. Outcomes could be either incidence of infections or change in hygiene behaviours. The Risk of Bias 2 tool was used to assess study quality.

**Results:** We found seven studies that met the inclusion criteria. Six studies reported successfully improving self-reported hygiene behaviour or health outcomes, but only one of these six trials confirmed improvements using objective measures (reduced consultations and antibiotic prescriptions), Germ Defence. Settings included kindergartens, workplaces, and service station restrooms. Modes of delivery were diverse: WeChat, website, text messages, audio messages to mobiles, electronic billboards, and electronic personal care records. Four interventions targeted parents of young children with educational materials. Two targeted the general population; these also used behaviour change techniques or theory to inform the intervention. Only one trial had low risk of bias, Germ Defence; the most common concerns were lack of information about the randomisation, possible bias in reporting of behavioural outcomes, and lack of an analysis plan and possible selective reporting of results.

**Conclusion:** There was only one intervention that was judged to be at low risk of bias, Germ Defence, which reduced incidence and severity of illness, as confirmed by objective measures. Further evaluation is required to determine the effectiveness of the other interventions reviewed.

## Introduction

COVID-19 is an infectious disease caused by a newly discovered coronavirus. The virus is spread primarily through droplets when an infected person coughs or sneezes, either travelling through the air directly to another person’s eyes, nose or mouth, or transmitted indirectly by landing on a surface, which is touched by a person who then touches their eyes, nose or mouth [1]. Therefore, we can prevent the spread of COVID-19 by adopting good hygiene practices, including hand washing, avoiding touching our faces, covering the mouth and nose when coughing or sneezing, and disinfecting surfaces [2]. Performing these behaviours can dramatically reduce the likelihood of household and community transmission [3].

In the absence of a vaccine, we are particularly reliant on behavioural measures and there is an urgent need to design interventions to improve hygiene practices [2]. Self-reported infection control behaviours during the pandemic are lower than is optimal for infection prevention [4]. Digital interventions are particularly desirable because of the ease of implementing them at scale and because they operate remotely, so can be rolled out without contact during the pandemic. We are the team that developed Germ Defence (https://germdefence.org/), a freely available website providing behavioural advice for infection control within households, using behaviour change techniques, which was shown to be effective at reducing the household transmission of infection during the H1N1 pandemic [5-7]. During the current COVID-19 pandemic, it is important to promote digital interventions for protective hygiene behaviours that have clear evidence of their effectiveness. We are re-purposing and disseminating Germ Defence and we wanted to investigate whether there are other interventions that we could learn from or that should be repurposed and promoted.

Therefore, we conducted a rapid systematic review to identify digital interventions that aimed to improve hygiene practices within the community. Our review question was: What is the evidence for the effectiveness of digital interventions used in household and community settings (e.g. schools, workplaces) for improving infection prevention and control behaviours and for reducing incidence of respiratory and gastro-intestinal infections?

## Methods

### Study design

The study protocol was registered on PROSPERO, registration number: CRD42020189919. We adopted best practice for rapid evidence reviews [8]. There were no deviations from protocol.

### Eligibility criteria

We used the following inclusion criteria:

i. Types of study to be included: only articles analysing or reporting data using the following designs: randomised controlled trials (including parallel, clustered, and crossover), controlled before-and-after trials, and interrupted time series analyses (with at least three timepoints before, and three timepoints after introduction of the intervention); this is because we were searching for studies that could provide high quality evidence of effectiveness of interventions.
ii. Condition being studied: Respiratory and gastro-intestinal infections, by which we mean infections that are spread by respiratory droplets and faeces, and hygiene interventions to prevent them; we deemed these to have similar infection prevention and control mechanisms to COVID-19.
iii. Participants/population: households and community settings (e.g. schools, workplaces).
iv. Intervention(s), exposure(s): Digital interventions aiming to improve hygiene, and infection prevention and control in the community. Digital interventions include those delivered via mobile messages, mobile apps, websites, social media, video games, virtual reality, remotely (e.g. via online coaching and networks), and wearable technology. Video recordings were only included if delivered via one of these methods. In any multi-component intervention that included non-digital elements, it must have been possible to extract data on the effectiveness of the digital component.
v. Comparator(s)/control: Any control group, could be either an active (non-digital) or a no-intervention control.
vi. Outcomes: health outcomes around transmission of infection and infection severity, or behavioural outcomes about increase in hygiene behaviours or quality of hygiene behaviours.

We used the following exclusion criteria:

i. Population: studies that are only in hospitals or that are delivered as a part of training for health care workers
ii. Interventions:
  a. interventions that are not concerned with hygiene measures to prevent transmission of respiratory and gastrointestinal infections, e.g., we excluded interventions focussing only on vaccine uptake, catheterisation or use of condoms.
  b. multi-component interventions that include non-digital elements, if it is not possible to extract data on the effectiveness of the digital component separately from the non-digital elements.
iii. Outcomes: studies reporting only outcomes on knowledge and attitudes, or intentions about behaviour, rather than behaviour.
iv. Study designs: Protocols, opinion pieces, commentaries, and interrupted time series with fewer than 3 timepoints before and 3 after introduction of the intervention.

We did not restrict the search by language, since we hoped to be able to extract data from relevant articles in any language. However, the databases that we searched required papers to have abstracts translated into either English or Chinese.

### Search strategy

We conducted a systematic literature search in five databases: MEDLINE, Embase, PsycINFO, Cochrane Controlled Register of Trials (CENTRAL), and China National Knowledge Infrastructure (CNKI). See Appendix 1 for the full list of search terms. The searches were conducted on 26^th^ May 2020. We restricted the searches to studies published since 1^st^ January 2000, since we did not expect that there would be any digital interventions prior to 2000, which is consistent to the approach taken by previous reviews [9].

We searched for relevant grey literature by checking key governmental websites (Center for Disease Control and Prevention, Public Health England, Chinese Center for Disease Control and Prevention, National Health Commission of the People’s Republic of China and National Administration of Traditional Chinese Medicine) and by checking reference lists of key relevant papers.

### Selection of studies

Titles and abstracts of studies in English were screened by two review authors (SD and LT) with disagreement resolved through discussion with a third (NG). Studies in Chinese were screened by RYX and XYH, there were no discrepancies. The full text of potentially eligible studies was independently assessed for eligibility by two review authors (NG and JG) and queries resolved by discussion with the full team.

The standardised Cochrane data collection form for intervention reviews was used to capture extracted data (https://dplp.cochrane.org/data-extraction-forms). One review author extracted data (in English LT for half the studies and SD for half the studies; in Chinese RYX) and this was double checked by a second author (in English JG, in Chinese XYH).

### Data synthesis

We planned to present a narrative synthesis of the results, and to conduct a meta-analysis if the interventions and outcomes measured were sufficiently similar.

### Risk of bias assessment

Risk of bias was assessed at the same time as data extraction using Risk of Bias 2.0 (RoB 2) [10]. Of interest for this review was the effect of the assignment to the intervention (the intention to treat (ITT) effect). We assessed the following types of bias as outlined in Chapter 8 of the Cochrane Handbook for Systematic Reviews of Interventions [11]: bias arising from the randomisation process, bias due to deviations from the intended interventions, bias due to missing outcome data, bias in measurement of the outcome, and bias in selection of the reported result. We used the signalling questions recommended in the tool to make a judgement regarding the likelihood of bias for each domain. Using the algorithms proposed by RoB 2, we then categorised each study according to the following levels of bias: low risk, some concerns, and high risk of bias. In order to be categorized as “low risk of bias”, a study had to be at low risk of bias for all domains. Studies were categorized as “some concerns” if they raised some concerns in at least one domain, but were not judged to be at high risk of bias for any domain. Finally, studies were categorized as “high risk of bias” if they were at high risk of bias in at least one domain, or were judged to have “some concerns” for multiple domains in a way that substantially lowered confidence in the results.

## Results

Searches of databases identified 3647 potentially relevant records. One additional article was identified through hand-searches and grey literature searches. There were a total of 3173 articles after duplicates were removed. After title and abstract screening, 70 articles were retrieved in full. In total, 7 studies met the inclusion criteria (Figure 1). A list of articles excluded after full-text screening is provided in Appendix 2.

**Figure 1:**
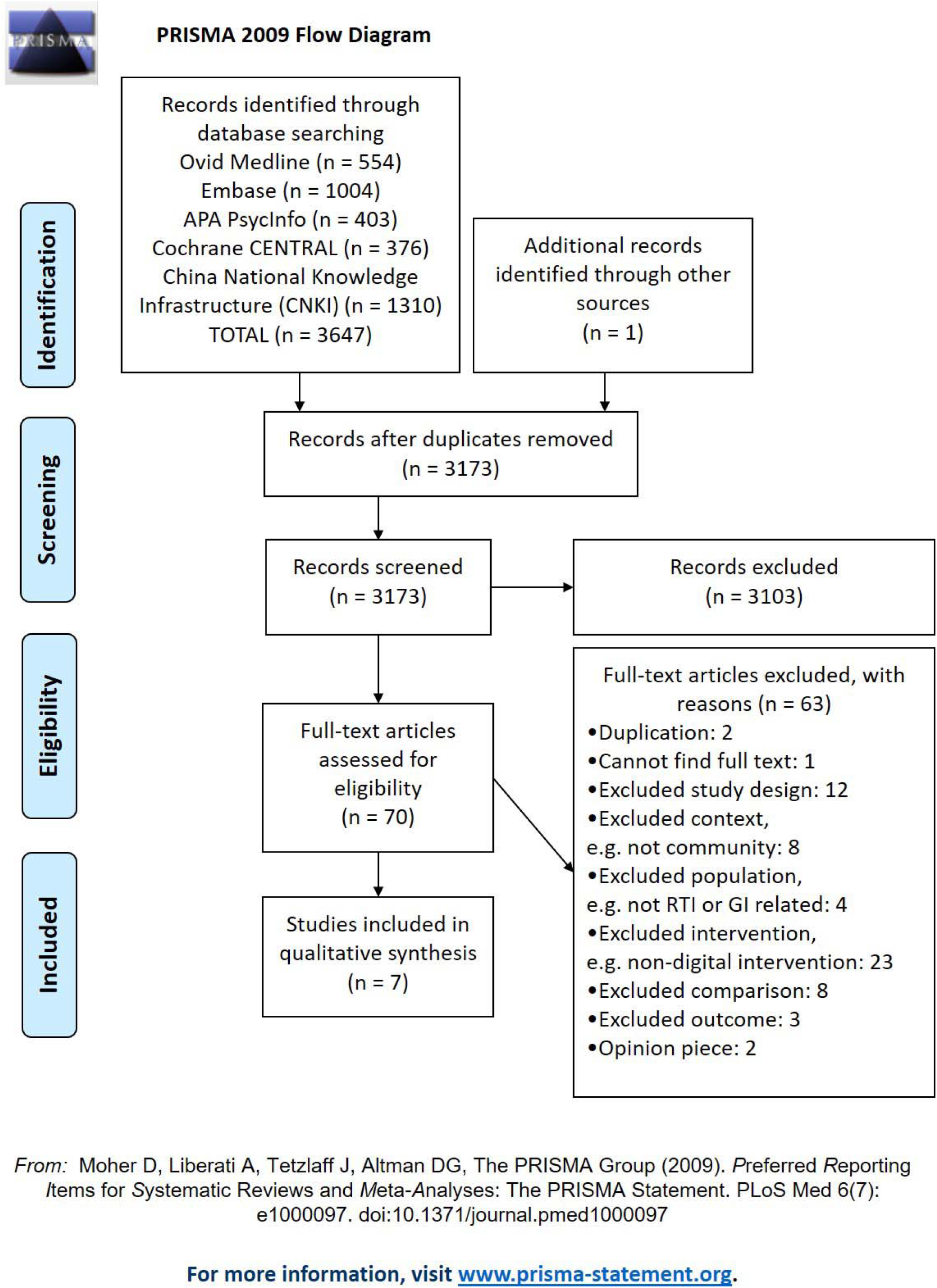
PRISMA flowchart

The studies were too heterogeneous for a meta-analysis, so we present a narrative synthesis of the results.

### Study Characteristics

We found seven studies that fulfilled our inclusion criteria [5, 12-17]. A summary of the study characteristics is in Table 1 and a detailed table of study characteristics is in Appendix 3. One of the papers, Tidwell 2019, reported two interventions, one of which was an excluded intervention—a TV advertisement—and is listed along with the excluded studies in Appendix 2. All were randomized controlled trials (RCTs), apart from Hu 2018, which was a nonrandomised but controlled trial [13]. One study reported only health outcomes (Hu 2019) [14], four reported only behavioural outcomes (Bourgeois 2008, Judah 2009, Tidwell 2019, and Wu 2020) [12, 15-17], and two reported both health and behavioural outcomes (Hu 2018, Little 2015) [5, 13].

**Table 1:** Summary study characteristics

| Study | Country | Design | Disease | Setting | Population | Mode of Delivery | Intervention(s) | Control | Duration of intervention | Outcome Measures |
| --- | --- | --- | --- | --- | --- | --- | --- | --- | --- | --- |
| Bourgeois 2008 | US | RCT | Flu | Workplace | 144 Healthy adults | Personally controlled health record program | Messages on influenza illness and prevention, tailored based on survey information provided and on postcode | Four monthly bulletins providing information on cardiovascular disease, stroke, skin cancer and sun protection, and guidelines for a healthy diet | 16 weeks | the rate of work attendance despite a respiratory illness, question in exit survey on hand hygiene question in exit survey on cough etiquette |
| Hu 2018 | China | Non-randomized but controlled trial | HFMD | Kindergartens and home | 10 staff, 60 parents and 60 healthy children | WeChat | WeChat education (group for GPs, staff, and parents) + Usual care | Usual care: health education, delivered face to face. | 3 months | Incidence of HFMD<br>Proportion of children who mastered the correct way of washing hands (measured at end of treatment)<br>Proportion of children who formed good habits of washing hands (measured at end of treatment) |
| Hu 2019 | China | RCT | HFMD | Community Health Service Center and home | Parents of 120 children with HFMD | WeChat | WeChat group for GPs and parents | Usual care: health education, delivered face to face. | 14 days | Duration of rash<br>Time to recovery |
| Judah 2009 | England | RCT | n/a | Service station bathrooms | Highway service station bathroom users, more than 198000 restroom uses over 32 days | Text-only message, displayed on an electronic dot matrix screen over the entryway to the 2 restrooms | 14 different intervention messages covering seven domains, max 48 characters, all included the word 'soap'. | Blank control: no message on the board<br>Positive control: "Wash your hands with soap." | July-September 2008 | Soap use ratio: Soap dispenser use divided by number of people in the bathroom during the intervention period (1-hour intervals) |
| Little 2015 | UK | RCT | RTIs | Online | 20,066 adult patients from 344 practices, who had at least one other individual living in the household | Web-based and email prompts | Four weekly web-based sessions, that provided information, developed a plan for handwashing, reinforced helpful attitudes and norms, addressed negative beliefs and used tailored feedback.<br>Automated emails were used to prompt participants to use sessions and complete questionnaires | No treatment: The control group did not have access to the intervention webpages, but they had monthly emails to prompt them to complete the questionnaires. | 4 months | Self-reported infections in user (incidence and severity of respiratory infections, incidence of gastrointestinal infections)<br>Self-reported incidence of respiratory infections in household members<br>Self-reported handwashing<br>Health care use as assessed from medical records (number of GP consultations and antibiotic prescriptions) |
| Tidwell 2019 Study 2 | India | RCT | n/a | Home | 617 new mothers, 605 mothers of 4-7 year olds | Mobile phone audio messages (all); reminder texts (new mothers only) | Audio phone messages of 90s, twice weekly, conveying health messages via a fictional dialogue between a doctor and a mother; Text reminders to practice the target behaviors | No intervention (though control sample also had to own a phone) | 8 weeks (new mothers), 4 weeks (mothers of 4-7 year-olds) | Handwashing with soap at key occasions: Measured 21 days after the end of the intervention using sticker diaries |
| Wu 2020 | China | RCT | HFMD | Home | 3000 parents of vaccinated healthy children aged 0-3 | Text messages | Messages of < 50 words, about knowledge, prevention and treatment of HFMD, sent at least weekly, with a total of 16-20 messages. | No intervention | 5 months | Proportion of children who wash hands before eating and after going to the toilet (measured at baseline and 1 year)<br>Proportion of children who wash hands after going out (measured at baseline and 1 year) |

Two of the studies were conducted in the UK (Little 2015, Judah 2009) [5, 15], one in the US (Bourgeois 2008) [12], three studies in China (Hu 2018, Hu 2019, Wu 2020) [13, 14, 17], and one in a relatively low income part of India (Tidwell 2019) [16]. All three Chinese studies concerned hand foot and mouth disease (HFMD) [13, 14, 17], one UK study focussed primarily on respiratory tract infections (RTIs, Little 2015) [5], and the one US study was on influenza (Bourgeois 2008) [12]. Two studies aimed to increase the incidence of hand washing with soap, without being focussed on the prevention of a particular disease (Judah 2009, Tidwell 2019) [15, 16].

Regarding the setting and population, three studies targeted parents of young children (Hu 2019, Tidwell 2019, Wu 2020) [14, 16, 17], one targeted parents and staff in kindergartens (Hu 2018) [13], one was set in the workplace (Bourgeois 2008) [12], one in service station restrooms [15], and one targeted the general adult population (Little 2015) [5]. The participants in the four studies that targeted parents of young children had a lower average age than participants in the two trials with a workplace or general population setting, see Appendix 3. The sample sizes varied from hundreds (Bourgeois 2008, Hu 2018 and Hu 2019) [12-14] to thousands (Tidwell 2019 and Wu 2020) [16, 17] to tens of thousands (Little 2015) [5] to hundreds of thousands (Judah 2009) [15]. One of the smaller trials, Bourgeois 2008, had a smaller sample than hoped for because it was set in the workplace and there was corporate restructuring during the recruitment period [12].

Two studies, Hu 2018 and Hu 2019, delivered their interventions via WeChat, a Chinese platform that is similar to Whatsapp, allowing users to send free messages to phone contacts that also use WeChat; to transfer pictures, videos, or speech; and enabling group chat [13, 14]. In both studies, a WeChat group was set up for health care workers and participants. Health care workers could upload health educational materials and participants could ask them questions about HFMD. Both WeChat interventions operated in the same manner, but they had different targets and therefore contents: one was for healthy children and included material that promoted knowledge of prevention methods (Hu 2018) [13], while one was for the parents of HFMD patients with a target of promoting knowledge of clinical treatment including nursing (Hu 2018) [14].

Germ Defence, Little 2015, is a web-based intervention plus email [5]. Participants were recruited from GP practices and offered a web-based session each week for four weeks, which provided information about the importance of influenza and the role of handwashing, encouraged users to develop a plan to maximise goal and intention formation for handwashing, reinforced helpful attitudes and norms, and addressed negative beliefs using tailored feedback. Automated emails were used to prompt participants (to complete the monthly questionnaires, and—in the intervention group—to use the sessions).

Tidwell 2019 sent weekly 90 second audio messages about hand hygiene to participants’ phones, supplemented with text message reminders to some participants [16]. The messages were branded as a part of a campaign and took the form of a fictional dialogue between a local doctor and a mother. Mothers of children from 4 to 7 years received messages for 4 weeks, whereas new mothers received messages for 8 weeks. Messages to the former were about hygiene topics, while messages to the latter included more general maternal health messages as well. New mothers also received reminder texts to practice the target behaviours.

Wu 2020 sent text messages of less than 50 words about knowledge, prevention and treatment of HFMD [17]. The messages were sent at least once a week starting one month before the peak time for hand foot and mouth, with a total of 16 to 20 messages over five months.

Bourgeois 2008 sent messages via a personally controlled health record program [12]. Enrolled subjects completed online health risk assessment surveys, the responses to which drove a decision support system to generate and send tailored health messages for participants in the intervention group. These messages were sent to participants’ personally controlled health record inbox, and participants were simultaneously notified with a standard, plain-text email instructing them to visit and log on to their personally controlled health record to review the message. The intervention group received messages on influenza illness and prevention, tailored based on information provided in baseline and seven bi-weekly surveys, and on postcode. There were five types of health message: vaccine reminders, respiratory illness advice, influenza alerts, weekly influenza risk maps, and monthly educational bulletins.

Judah 2009 used electronic billboards at the entrance to service station restrooms [15]. Messages were a maximum of 48 characters, were in capital letters, and flashed for the duration of their presentation to attract attention. The researchers tested two messages from each of seven domains; all included the word ‘soap’. The domains were:

- Knowledge of risk: Inform people about a fact they may not know;
- Knowledge activation: Remind people of what they know already or convince them of the importance of what they know;
- Norms or affiliation: Raise concern for social judgments on people’s hygiene behaviours because of the knowledge that others might be concerned with standards for acceptable behaviour;
- Status or identity: Help people to feel that hand washing—or more broadly, cleanliness and being hygienic—is an important aspect of their self-image;
- Comfort. Emphasize positive sensory qualities of having clean hands;
- Disgust: Trigger the arousal of a ‘ ‘yuck’ ‘ response;
- Cue. Provide people with a behavioural rule triggered by an object in the environment or an event.

The study using the personally controlled health record program (Bourgeois 2008) had an active control condition [12]. Control participants were sent monthly bulletins (four in total) providing information on cardiovascular disease, stroke, skin cancer and sun protection, and guidelines for a healthy diet. The service station restroom trial had both a passive control (blank board) and an active control (“Wash your hands with soap”). The other trials all had a no intervention/ usual-care control.

In three studies, Judah 2009, Tidwell 2019 and Wu 2020, participants were passive receivers of the intervention [15-17]. In two, Little 2015 and Bourgeois 2008, studies they completed questionnaires [5, 12]. And in two studies, Hu 2018 and Hu 2019, the participants could interact with each other and with health care professionals [13, 14].

All seven studies either sent participants educational material or, in the case of Judah 2009 [15], had conditions that targeted knowledge. Both of the studies that asked participants complete questionnaires (Little 2015 and Bourgeois 2008) also used them to provide tailored feedback [5, 12]. One of these two studies, Little 2015, also used planning to support the formation of intentions to wash hands, monitored handwashing behaviour, reinforced helpful attitudes and norms, and addressed negative beliefs [5]. Three studies sent prompts as well as the main intervention: Bourgeois 2008 sent emails to remind the participants to log-on to their personal health care record [12], Little 2015 to remind them to complete questionnaires and use the intervention sessions [5], and Tidwell 2019 sent texts to one group of participants to remind them to practice the target behaviours [16].

The restroom trial messages in Judah 2009 were developed at a workshop of experts, based on empirical data and theoretical domains from behaviour change theory [15]. Tidwell 2019 piloted messages on a group that was similar to the target participants, to check for comprehensibility and acceptability of content [16]. Little 2015 developed the Germ Defence intervention iteratively with users [5].

### Risk of bias assessment

The risk of bias assessment for each study is reported in Table 2. There was one trial at low risk of bias, Little 2015 [5], four where there were some concerns, Bourgeois 2008, Hu 2019, Tidwell 2019 and Wu 2020 [12, 14, 16, 17] and two with high risk of bias, Hu 2018 and Judah 2009 [13,15]. The risk of bias ratings are summarized in Table 2.

- Bias due to randomisation process: four of the trials lacked information about the randomisation process (Hu 2019, Judah 2009, Tidwell 2019 and Wu 2020) [14-17], which caused some concerns. A fifth, Hu 2018, did not employ randomisation between groups, so it has a high risk of bias [13].
- Bias from deviation from intended interventions: in three of the studies, Bourgeois 2008, Hu 2019 and Tidwell 2015, participants were aware which trial arm they had been assigned to [12, 14, 16], which raises some concerns; in one cluster RCT, Bourgeois 2008, the participants were aware that they were in a trial and trial personnel were aware of participants’ assignments to conditions [12], which the tool rates as giving some concerns—in both cases, the idea being that when participants or trial personnel know their assignment then this may lead to deviation from intended interventions
- Bias due to missing outcome data: there were some concerns over missing outcome data in one trial where there was a technical malfunction, Judah 2009 [15], and in one trial where there was a lack of information, Tidwell 2019 [16].
- Bias due to measurement of the outcome: two of the studies had primary outcomes that were self-reported (Bourgeois 2008 and Tidwell 2019), which caused some concerns [12, 16], and three studies (Hu 2018, Hu 2019 and Wu 2020) had no information on who assessed the outcome measure, which caused some concerns [13, 14, 17].
- Bias due to selection of the reported result: three studies (Hu 2018, Hu 2019 and Wu 2020) lacked both information about an analysis plan and enough information to know if they may have selectively reported outcome measures, which caused some concerns [13, 14, 17]; one study (Judah 2009) [15] lacked information about an analysis plan and had multiple outcomes that were selectively reported (the results were split by gender and the blank passive control was separated from the positive active control, in neither case were the pooled results reported), which puts it as high risk of bias.

**Table 2:** Risk of Bias 2.0 Quality Assessment

| Study | Bias due to the randomisation process | Bias due to deviations from the intended interventions | Bias due to missing outcome data | Bias in measurement of the outcome | Bias in selection of the reported result | Overall risk of bias |
| --- | --- | --- | --- | --- | --- | --- |
| Bourgeois 2008 | Low | Some concerns | Low | Some concerns | Low | Some concerns |
| Judah 2009 | Some concerns | Low | Some concerns | Low | High | High |
| Hu 2018 | High | Some concerns | Low | Some concerns | Some concerns | High |
| Hu 2019 | Some concerns | Some concerns | Low | Some concerns | Some concerns | Some concerns |
| Little 2015 | Low | Low | Low | Low | Low | Low |
| Tidwell 2019 | Some concerns | Some concerns | Some concerns | Some concerns | Low | Some concerns |
| Wu 2020 | Some concerns | Low | Low | Some concerns | Some concerns | Some concerns |

### Health Outcomes

Three trials reported health outcomes, Little 2015, Hu 2018 and Hu 2019 [5, 13, 14]. Little 2015 was conducted in the UK, focussed on RTIs, and was judged to be at low risk of bias; Hu 2018 and Hu 2019 were conducted in China on HFMD; Hu 2018 was judged to be at high risk of bias and Hu 2019 was judged have some concerns. All reported that they were successful at reducing either incidence or duration of illness, and Little 2015 was also successful at reducing the use of healthcare resources (consultations and antibiotics). See Table 3.

**Table 3:** Summary of health outcomes

| Study | Outcome Measure | Outcome in Control Group | Outcome in Intervention Group | Test statistics and statistical significance (where available) |
| --- | --- | --- | --- | --- |
| Hu 2018 | Incidence of HFMD | 13.3% (4/30) | 0% (0/30) | n/a |
| Hu 2019 | Duration of rash in days, M (SD) | 7.43 (1.9) | 3.65 (0.8) | $p < 0.05$ |
| | Time to recovery in days, M (SD) | 13.04 (2.6) | 6.66 (1.5) | $p < 0.05$ |
| Little 2015 | Number reporting one or more episodes of RTI at 16 weeks | 59% (5135/ 8667) | 51% (4242/ 8241) | Multivariate risk ratio 0.86; 95% CI 0.83–0.89; $p < 0.0001$ |
| | Number reporting one or more episodes of gastrointestinal infections at 16 weeks | 25% (1821/ 7292) | 21% (1376/ 6410) | Multivariate risk ratio 0.82; 95% CI 0.76-0.88; $p < 0.0001$ |
| | Number of respiratory infections at 4 months, M (SD) | 1.09 (1.36) | 0.84 (1.13) | Multivariate incident rate ratio 0.75; 95% CI 0.72-0.79; $p < 0.0001$ |
| | Number of days of moderate or bad symptoms, M (SD) | 2.60 (4.44) | 2.08 (4.00) | Multivariate incident rate ratio 0.92; 95% CI 0.87-0.98; $p < 0.0001$ |
| | Antibiotic use in primary care within 4 months | 6% (617/9579) | 6% (535/9540) | Multivariate risk ratio 0.83; 95% CI 0.74-0.94; $p = 0.002$ |
| | Antibiotic use in primary care within 12 months | 11% (1008/9579) | 9% (891/9540) | Multivariate risk ratio 0.85; 95% CI 0.77-0.93; $p < 0.001$ |
| | Consultation in primary care or hospitalisation with respiratory infection within 4 months | 11% (1021/9579) | 10% (951/9540) | Multivariate risk ratio 0.90; 95% CI 0.82-0.98; $p = 0.014$ |
| | Consultation in primary care or hospitalisation with respiratory infection within 12 months | 17% (1653/9579) | 16% (1527/9540) | Multivariate risk ratio 0.90; 95% CI 0.84-0.96; $p = 0.001$ |
| | Number of respiratory infections in household members | 49% (4193/ 8551) | 44% (3545/ 8075) | Multivariate risk ratio 0.88; 95% CI 0.85-0.92; $p < 0.0001$ |
| | Number of respiratory infections in the household at 4 months, M (SD) | 1.17 (2.07) | 0.93 (1.48) | Multivariate incident rate ratio 0.79; 95% CI 0.74-0.83; $p < 0.0001$ |

The Germ Defence intervention was successful at reducing incidence of RTI and transmission within the home [5]. After 16 weeks, 51% of individuals in the intervention group reported one or more episodes of RTI compared with 59% in the control group (multivariate risk ratio 0-86, 95% CI 0-83—0-89; p<0-0001). The intervention reduced transmission of RTIs (reported within 1 week of another household member) both to and from the index person. It also reduced incidence of gastrointestinal infections from 25 % in the control to 21% in the intervention (multivariate risk ratio 0.82, 95% CI 0.76-0.88; p<0.0001). The participants in the intervention group had less days of moderate or severe symptoms. These self-report measures were confirmed by objective measures from medical notes: the intervention group were less likely to have had antibiotics prescribed, or to have had a consultation in primary care or hospitalisation with RTI, at both four and twelve months.

Two Chinese studies, Hu 2018 and Hu 2019, one which was at high risk of bias and one that had some concerns, reported that establishing a WeChat group could improve health outcomes related to HFMD [13, 14]. Hu 2019 was an RCT, and reported that the WeChat intervention decreased the duration of the rash and led to faster recovery compared to a control group that only received the usual face-to-face GP care: duration of rash: intervention *M (SD)=* 3.65 (0.8) days, control *M (SD)=* 7.43 (1.9) days; time to recovery: intervention *M (SD)=* 6.66 (1.5) days, control *M (SD)=* 13.04 (2.6) days [14]. Hu 2018 was a nonrandomised but controlled trial, where a WeChat was established among parents and staff in a daycare setting. They reported that the incidence of HFMD decreased from 13.3% in the control to 0 in the group that had the WeChat intervention [13].

### Behavioural Outcomes

Six studies reported behavioural outcomes: Bourgeois 2008, Hu 2018, Judah 2009, Little 2015, Tidwell 2019 and Wu 2020 [5, 12, 13, 15-17]. They were all concerned with handwashing. Three trials measured hand washing via self-report, Bourgeois 2008, Little 2015 and Tidwell 2019 [5, 12, 16], whereas two reported behaviour about handwashing without saying who took the measure (Hu 2018 and Wu 2020) [13, 17], and one reported soap use ratio (amount of soap used divided by number of restroom users), Judah 2009 [15]. Five out of the six (all but Bourgeois 2008) reported that the interventions increased handwashing (or soap use as a proxy for handwashing) [5, 13, 15-17]. (See Table 4.) Judah 2009 and Hu 2018 were judged to be at high risk of bias, and there were some concerns about Bourgeois 2008, Tidwell 2019 and Wu 2020.

**Table 4:**
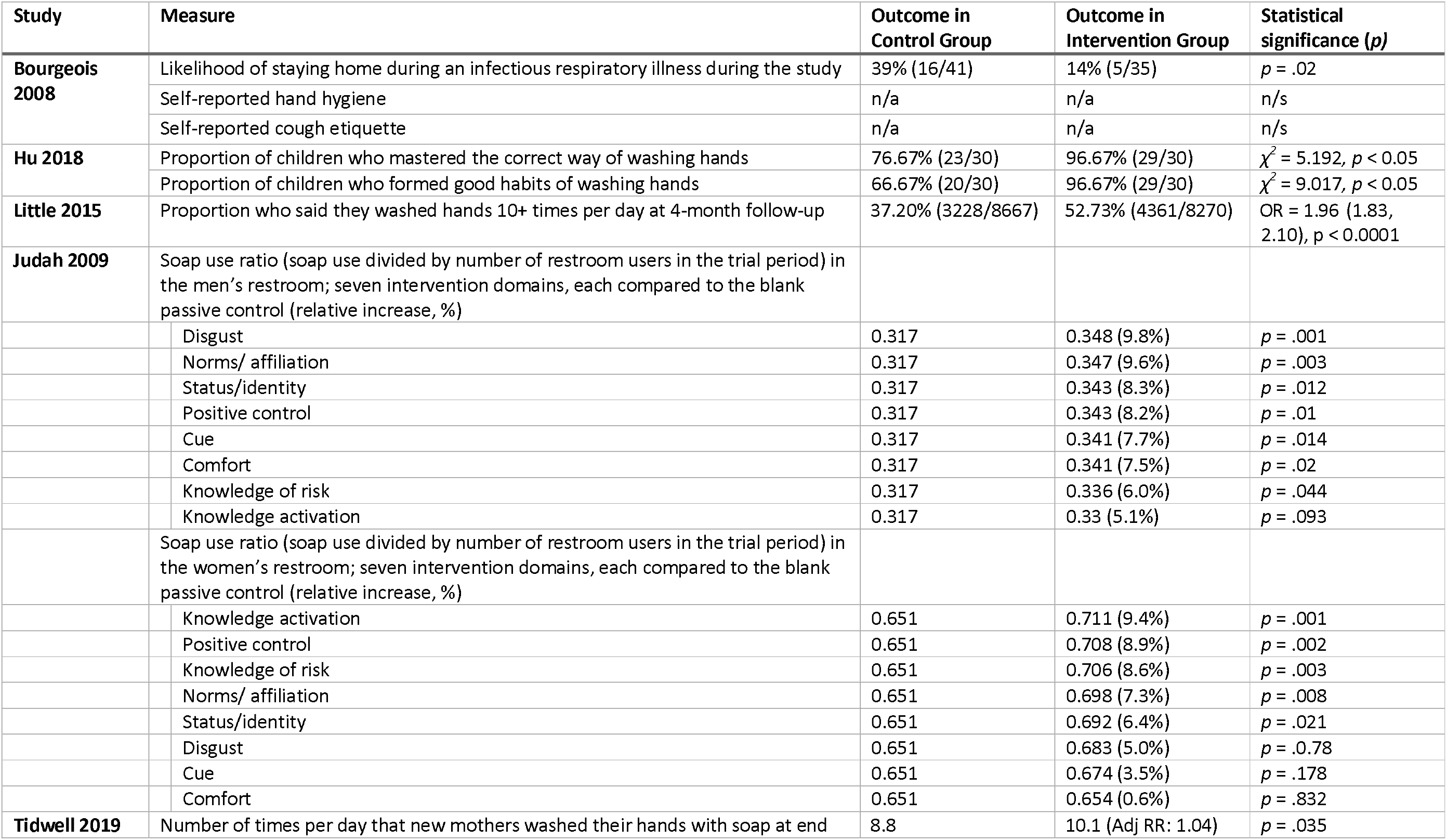
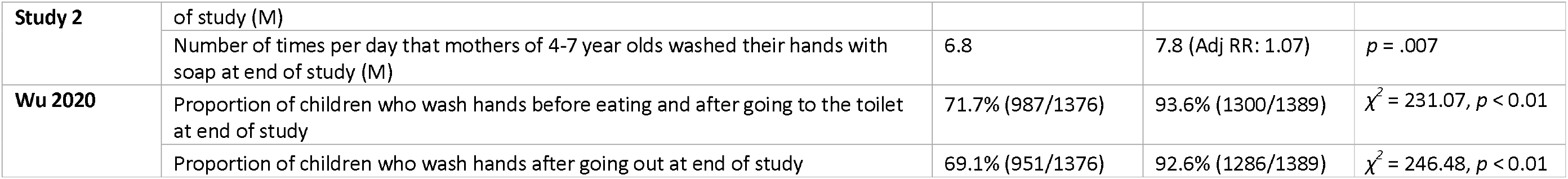
Summary of behavioural outcomes

Wu 2020 found that sending weekly text messages about prevention and treatment of HFMD to parents increased handwashing among children [17]. It is not clear from the paper who took observations for the outcome measure or whether they were self-reported. One year after the start of the trial the proportion of children in the intervention group who washed their hands after going out was 92.6%, compared to 68.1% in the control, and the proportion who washed their hands before and after eating one year after the start of the intervention was 93.6% in the intervention group compared to 71.7% in the control (both *p* < 0.01).

Sending weekly 90 second audio messages about hand hygiene to mothers in a relatively low-income area of India led to increased self-reported handwashing with soap at key occasions [16]. Hand washing behaviour was measured 21 days after the end of the intervention, by putting stickers to represent handwashing occasions in a sticker diary, which had the day segmented into 7 sections; participants also had to self-report some distractor activities, which were intended to mask the purpose of the study. There were two groups in the study, new mothers and mothers of 4 to 7-year olds, each with a comparable control group (who also had to own phones). New mothers were more likely to report washing their hands in the intervention than in the control group (Adj RR: 1.04, *p* = .035), corresponding to 1.3 more occasions daily, and a 3.0 percentage point increase from a baseline rate of 49.6%. The mothers of 4 to 7-year olds were more likely to report washing their hands in the intervention group than the controls (RR: 1.07, *p* = .007), corresponding to 1.0 more occasions daily, and an increase of 3.4 percentage points over a baseline rate of 46.7%. The base rates were 8.8 self-reported handwashes with soap at key times in the new mothers intervention and 6.8 for mothers of 4 to 7-year olds, so this corresponds to an increase to a mean of 10.1 self-reported handwashes per day for new mothers and 7.8 for mothers, of 4 to 7-year olds, a 15% relative increase in each group.

In Hu 2018’s non-randomized but controlled trial using an interactive WeChat intervention, where health care workers could circulate educational materials and parents could ask questions, both the proportion of children who mastered the correct way of washing their hands and the proportion of children who formed good habits of washing their hands increased from 76.7% to 96.7% (both *p* < 0.05) [13]. It is not reported how the measurement were taken, or whether they were self-reports.

Putting electronic text-only messages on bulletin boards outside of English highway service station restrooms was effective at increasing soap use ratio (soap use/ per person entering the restroom) [17]. Most of the seven intervention domains in Judah 2009 showed a small but statistically significant increase in soap-use ratio when compared with the blank passive control; however, the pattern of results was very different for men and women. Knowledge activation (reminders about the dangers of failing to wash hands) was the top-performing domain for women with a 9.4% increase compared with the blank control condition, *p* = .001, but was ineffective for men. Disgust messages, which aimed to arouse a “yuck” response, led to the biggest improvement in men with a 9.8% relative increase compared with the blank control, *p* = .001, but produced no significant response in women. Norms and status/ identity were effective for both genders, as was the positive control condition, “Wash your hands with soap”. There were two messages for each domain and the only individual message that was effective for both genders was the norms message, ‘ ‘Is the person next to you washing with soap?’’, which resulted in a 12.1% relative increase in hand-washing ratio among men and a 10.9% increase among women compared with the control condition. This was the most effective message for men and the second most effective for women.

A trial in a US workplace, Bourgeois 2018, that sent tailored targeted messages about influenza illness and prevention over 16 weeks, via a personally controlled health record program, had mixed results, including no statistically significant improvement in handwashing [12]. Participants in the intervention group were more likely to stay home during an infectious respiratory illness (self-reported) compared with participants in the control group, who received messages about cardiovascular care and sun protection (39% [16/41] vs 14% [5/35], respectively; *p* = .02) There was no change in self-reported hand hygiene or cough etiquette.

Little 2015 found that self-reported handwashing was higher in the group that had received the Germ Defence intervention [5]. The proportion of participants who reported that they washed their hands 10+ times per day was 52.7% in the intervention group, compared to 37.2% in the control, OR = 1.96 (1.83, 2.10), *p* < 0.0001.

## Discussion

### Principal findings

This rapid review synthesised evidence from controlled trials on the effectiveness of digital interventions to improve hygiene practices for infection prevention in the community, evaluated by improvements in health or behavioural outcomes. We found six randomized controlled trials and one non-randomized but controlled trial. Only one trial (of Germ Defence) provided good evidence of effectiveness, based on reduced reported incidence and severity of illness, confirmed by objective measures of reduced consultations and antibiotic prescriptions [5]. No other trials were low risk of bias or had objective evidence of effectiveness for health outcomes, and so further evaluation is required to determine the effectiveness of the other interventions reviewed.

All studies had a core educational message or, in the case of the restroom trial of Judah 2009 [15], two of the domains tested involved knowledge (either giving people new facts or activating knowledge that people would already be expected to have). Two studies also used behaviour change techniques (Little 2015’s Germ Defence [5]) or were based on behaviour change theory (Judah 2009’s billboards outside restrooms [15]). The trial of billboards outside restrooms found that knowledge activation (reminding people of something they already know) worked well for increasing the soap use ratio of women but not men, and disgust (triggering a yuck response) worked for well men but not women [15]. (Two different messages for each domain.) The norms message, ‘‘Is the person next to you washing with soap?’’, was effective in both groups, being the single most effective message for men and the second most effective for women. The simple reminder, “Wash your hands with soap”, was also effective for both groups.

The modes of delivery included websites, text messages, audio messages, WeChat, personal electronic health records, and electronic billboards outside restrooms. Three of the trials supplemented the main content and mode of delivery with either email or text message reminders. There is no indication that this extra input increased the effect size. In two cases, Bourgeois 2008 and Little 2015, emails were sent to participants as prompts to engage with the intervention (go to the website or log-on to their personal healthcare record) [5, 12]. In one case, Tidwell 2015 [16], the content of the text was a reminder to practice behaviours. However, this trial contained two groups, new mothers and mothers of 4 to 7-year olds, and only the new mothers received texts but both groups saw a 15% relative increase in self-reported handwashing.

Only one trial with behavioural outcomes did not find that the intervention improved the measure of hand washing, Bourgeois 2018 [12]. The studies were so heterogeneous that there could be many reasons for this: the workplace setting, the delivery of the messages by personal health care record, the US setting, or the content of the messages.

Of the six studies that reported successfully improving hygiene behaviour or health outcomes, four were targeted specifically at parents of young children (Hu 2018, Hu 2019, Tidwell 2019, and Wu 2020) [13, 14, 16, 17]. Only two targeted the general population (Little 2015 and Judah 2009) [5, 15]. Of these, Judah 2009 [15] demonstrated an increased soap ratio among the general British travelling public but, because of the nature of the trial, there is no precise demographic information about the participants. Germ Defence demonstrated reduced transmission of infection in a wide range of households, including those with comorbid illness, which is a particularly important at-risk group for COVID-19 [5].

Three of the studies had smaller sample sizes [12-14]—in the hundreds, as compared to the thousands to hundreds of thousands in the other four studies. In one case (Bourgeois 2019), this was because there was a workplace setting and corporate restructuring during the recruitment period led to a smaller sample than hoped for [12]. The other two studies used WeChat (Hu 2018, Hu 2019) [13, 14]. This is a Chinese platform that is similar to Whatsapp and, in the studies, GPs could upload educational materials to the group and answer questions from participants. This active involvement of GPs may be a reason why sample size was smaller than some of the other interventions and might make the intervention less scaleable for a large population.

### Strengths and limitations

We were able to search a wide range of papers because we did not have a language restriction and we specifically searched Chinese-language repositories, finding three Chinese-language papers. However, because we conducted a rapid review we searched a limited number of databases, so we may have missed papers. In particular, we may have missed papers that were in languages other than English or Chinese. Our search terms were in English and Chinese, and the English-language databases would only have recorded foreign-language papers if their abstracts had been translated into English.

Other major limitations relate to the studies themselves. The interventions and outcome measures were heterogeneous, which precluded a meta-analysis. Only one was deemed to be at low risk of bias, Little 2015 [5]. The most common faults were that the studies lacked information about the randomisation process, that they did not give information about an analysis plan and had the potential for selective reporting of results, and that there was risk of bias in the measurement of outcomes in behavioural studies. In many of the trials either participants or personnel were aware of the assignment to conditions, which the RoB 2 tool rates as being at some concern of bias from deviation from intended interventions. Arguably, being aware of the assignment is not a source of risk in digital interventions, especially those where the intervention is completely fixed in advance, so participants and personnel have no influence over delivery of the intervention.

### Implications for policy and practice

We limited our search to studies that aimed to increase community hygiene practices or to decrease infections that are transmitted by respiratory droplets or from faeces, so that the mode of transmission is similar to SARS-CoV-2. Therefore, the interventions we found could be relevant for preventing infection transmission in the current COVID-19 pandemic.

Germ Defence, which was trialled in the H1N1 pandemic and for seasonal flu in subsequent years, demonstrated reduced transmission in a wide age range of UK households, including ‘at risk’ individuals with chronic health conditions [5]. This is underpinned by an increase in self-reported handwashing (see also [6]) and supported by objective data from medical notes. It has already been re-purposed for COVID-19 and disseminated in the UK, China, and other countries [4].

A second intervention that may be worth re-purposing put messages on electronic billboards at the entrance to service station restrooms [15]. This would likely be cost-effective and scaleable, though the trial was judged as being at high risk of bias. It was trialled in England and, if implemented in a different country, one would need to consider the applicability of messages across cultures, though the top-performing social norms message and the reminder to wash hands with soap seem likely to have universal appeal. However, it should be noted there were 16 messages (including the two controls) shown in one-hour blocks in a motorway service station. Most participants would not have used the restrooms regularly or been exposed to the same message multiple times. If the same message were shown in front of all restrooms, people might become habituated and the effect of the message might decline over time. Therefore, repurposing this intervention would need to involve further refinement to show that habituation could be prevented, maybe by having a revolving suite of messages.

Of the four other effective interventions, all targeted parents of young children with educational materials, three about HFMD and one about handwashing for children in a relatively low-income part of India. All four were mainly educational. Three had some concerns about risk of bias and one was judged to be high risk. Before implementing them for COVID-19, there would need to be careful consideration of their applicability across diseases and—if being introduced in a different country from where they were tested—across cultures. Two of these used a WeChat intervention [13, 14], which might not be scaleable because it involved interaction with health care workers.

### Conclusions

Evidence from controlled trials shows that digital interventions can be effective at improving hygiene behaviours in the community, especially handwashing; and at decreasing the incidence, transmission, and severity of infections. There was one intervention with a low risk of bias that showed a reduced risk of infection: Germ Defence. It has the potential to be repurposed and used in the COVID-19 pandemic.

## Data Availability

All relevat data is available within supplementary materials, and any further data can be requested from the authors. All relevant data will be hosted in a Figshare repository upon final acceptance of the paper.

## Registration

PROSPERO CRD42020189919.

